# Generalized and specific whole-brain white matter abnormalities in human cocaine and heroin use disorders

**DOI:** 10.1101/2022.03.22.22272765

**Authors:** Pierre-Olivier Gaudreault, Sarah King, Pias Malaker, Nelly Alia-Klein, Rita Z. Goldstein

**Author notes:** **Corresponding author:** Rita Z. Goldstein, PhD, Mount Sinai Professor in Neuroimaging of Addiction, Department of Psychiatry (primary) and Department of Neuroscience (secondary), Chief, Neuropsychoimaging of Addiction and Related Conditions (NARC) Research Program, Icahn School of Medicine, Gustave L. Levy Place, Box 1230, New York, NY 10029.

## Abstract

Neuroimaging studies in substance use disorder have shown widespread impairments in white matter (WM) microstructure suggesting demyelination and axonal damage. However, substantially fewer studies explored generalized vs. acute/specific drug effects on WM. Our study assessed whole-brain WM integrity in three subgroups of addicted individuals, encompassing cocaine (CUD) and heroin (HUD) use disorder, compared to healthy controls (CTL).

Diffusion MRI was acquired in 58 CTL, 28 current cocaine users/CUD+, 32 abstinent cocaine users/CUD−, and 30 individuals with HUD (positive urine for cocaine in CUD+ and opiates used for treatment in HUD). Tract-Based Spatial Statistics allowed voxelwise analyses of diffusion metrics [fractional anisotropy/FA, mean diffusivity/MD, radial diffusivity/RD, and axial diffusivity/AD]. Permutation statistics (p-corrected<.05) were used for between-group t-tests.

Compared to CTL, all drug-addicted individuals showed widespread FA reductions, and increased MD, RD, and AD (19-57% of WM skeleton, p<.05). The HUD showed the most impairments, followed by the CUD+, with only minor FA reductions in CUD− (<.2% of skeleton, p=.05). Longer periods of regular use were associated with decreased FA and AD, and higher subjective craving was associated with increased MD, RD, and AD, across all drug-addicted individuals (p<.05).

These findings demonstrate extensive WM impairments in drug-addicted individuals characterized by decreased anisotropy and increased diffusivity, thought to reflect demyelination and lower axonal packing. Extensive abnormalities in both groups with positive urine status (CUD+ and HUD), and correlations with craving, suggest greater WM impairments with recency of use. Results in CUD−, and correlations with regular use, further imply cumulative and/or persistent WM damage.

## Introduction

Drug addiction is a complex and chronic brain disorder that encompasses periods of intoxication and compulsive drug-seeking usually followed by withdrawal and drug-related cravings that repeat despite adverse consequences (1). Functional neuroimaging studies have consistently revealed cognitive and emotional impairments in individuals with substance use disorders (SUD), associated with the recruitment of multiple large-scale brain networks (including reward, habit, salience, executive, memory, and self-directed networks) during drug-related processing, and reduced responses in these networks during non-drug-related processing [see (2) for an extensive review]. The neurobiological correlates of these dysfunctions implicate gray and white matter deficits, especially in the prefrontal cortex (PFC) but also in tracts projecting to and from the PFC [see (3) for a recent review].

Using diffusion tensor imaging (DTI), the current state-or-the-art technique to quantitatively assess the brain’s white matter (WM) microarchitecture, several studies reported impairments in nicotine, alcohol, cannabis, methamphetamine, cocaine, and opiate use disorders as recently reported in a review (4), a meta-analysis (5), and a mega-analysis (6). In individuals with addictions to cocaine and heroin, the two illicit drugs considered to cause the most severe dependence and harm (7), WM abnormalities have been observed in all major WM tracts, especially in projections to and from PFC regions. These included decreased fractional anisotropy (FA) and increased mean diffusivity (MD) in cocaine addiction (8–11), suggestive of myelin damage. Higher radial (RD) and axial diffusivities (AD), together with reduced FA, suggest also alterations in axonal integrity as observed in heroin-addicted individuals (12–16). Furthermore, impaired WM microstructure in the tapetum in cocaine users (10), and more globally in the frontal WM of heroin users (14,17), was correlated with longer duration of drug use, suggesting the impact of the chronicity of use.

However, there has been relatively less emphasis on studying the impact of addiction generally vs. acute/specific effects of a particular drug on these WM microstructural alterations. To the best of our knowledge, only one study directly compared stimulant- and opiate-addicted individuals, showing frontal WM hyperintensities in T2-weighted MRI images in both groups compared to healthy subjects (18). In this prior study, compared to opiate users, cocaine users showed both higher prevalence (including global, deep brain, and insular hyperintensities) and severity of these abnormalities, which may reflect the more severe cardiovascular effects associated with cocaine use (19). In addition, stimulants directly bind to the dopamine transporters (20) whereas opiates act indirectly on the dopaminergic system by activating the mu opioid receptors on GABAergic interneurons in the ventral tegmental area (21). Given that the dopaminergic system modulates axonal myelination throughout the brain (22–24), such variability in their effects on meso-striatal dopamine could serve as another basis for postulating a differential pattern in WM abnormalities between these drug classes.

The objective of this whole-brain diffusion MRI study was to compare healthy controls and individuals with SUD, including two cohorts of individuals with cocaine use disorder (CUD) or heroin use disorder (HUD). We further explored individual differences within the CUD group by including two subgroups [abstinent = negative urine toxicology for cocaine (CUD−) vs. current users = positive urine toxicology for cocaine (CUD+)] to test the potential impact of acute drug effects. Based on prior studies, we expected to observe WM abnormalities, characterized by reduced FA and increased MD, RD, and AD, across all individuals with SUD when compared to healthy controls. Given the mean length of abstinence in the CUD−group (>1 year), and in accordance with our previous study (25), we expected less pronounced WM abnormalities in this subgroup as compared with the CUD+ group. Finally, a direct comparison between the HUD group and both CUD subgroups was intended to further our understanding of the role of general addiction (to cocaine or heroin) vs. specific drug effects on WM. Given the positive urine status of the HUD group (on medically-assisted treatment (MAT) and hence putatively more similar to the CUD+) but also their time since last heroin use (>6 months and hence putatively more similar to the CUD−), directionality of these effects was exploratory.

## Methods and Materials

### Participants

Ninety individuals with SUD (60 with CUD and 30 with HUD undergoing MAT) and 58 healthy controls were recruited as part of two research protocols [data for 27 healthy controls and 25 CUD was previously reported (6)]. Subjects with CUD were recruited by advertisements and flyers as well as from educational talks provided at collaborating substance abuse treatment institutes in the New York metropolitan area. The CUD group included 32 abstinent users (average abstinence: 18 months) and 28 current users (average abstinence: 3 days). Healthy controls were recruited from the same communities for matching purposes. Individuals with HUD were recruited from a single inpatient drug addiction rehabilitation facility (Samaritan Daytop Village, NY). All participants provided written informed consent and study procedures were approved by the Icahn School of Medicine at Mount Sinai’s institutional review board. All SUD subjects met criteria as assessed by the Structured Clinical Interview for the *Diagnostic and Statistical Manual of Mental Disorders, fourth or fifth editions* (26,27) (for the CUD) or the Mini International Neuropsychiatric Interview (28) (for the HUD), and the Addiction Severity Index (29) for all subjects. The CUD participants met criteria for current cocaine dependence (n=33), abuse (n=3), or dependence in remission (n=24). All HUD participants met criteria for SUD with heroin being their primary drug of choice. Within the CUD sample, nine also met criteria for HUD, and within the HUD sample, nine also met criteria for CUD; however, these individuals’ primary drug of abuse was determined to be congruent with their assignment to their respective groups. The route of drug administration included smoking (41 CUD/3 HUD), intra-nasal (18 CUD/11 HUD), intravenous (1 CUD/15 HUD), and oral (1 HUD). Other comorbidities included alcohol use disorder (21 CUD/4 HUD), marijuana use disorder (9 CUD/2 HUD), amphetamine use disorder (1 CUD/3 HUD), and post-traumatic stress disorder (1 CUD/1 HUD). All SUD comorbidities were in partial or sustained remission at the time of study. For the CUD or HUD groups, respectively, symptoms of withdrawal were assessed with the Cocaine Selective Severity Assessment (30) or the Subjective Opiate Withdrawal Scale (31); and symptoms of craving were assessed with the 5-item Cocaine Craving Questionnaire (32) or the Heroin Craving Questionnaire – Short form-14 (33) on the day of the scan. These scores were range-corrected to a common scale for group comparisons. Dependence severity was assessed with the Severity of Dependence Scale (34) and the Fagerstrom Test for Nicotine Dependence (35) was used to measure nicotine dependence in all subjects. Recency of drug use was assessed in all participants objectively with a urine toxicology test. Urine was positive for cocaine for the CUD+; it was negative for the CUD−. With the exception of one participant, all individuals with HUD were urine negative for heroin, but all tested positive for other opiates [those used for MAT: methadone n=25 (106.5±62.0mg, 1 missing), buprenorphine/naloxone n=5 (14.7±8.3mg, 2 missing)]. Exclusion criteria were: 1) present or past history of DSM-IV or DSM-5 diagnoses of psychotic disorder or neurodevelopmental disorder; 2) history of head trauma with loss of consciousness (>30 min); 3) history of neurological disorders including seizures; 4) current use of any medication (with the exception of MAT in the HUD) that may affect neurological functions; 5) current medical illness and/or evident infection including cardiovascular disease (e.g., high blood pressure), as well as metabolic, endocrinological, oncological or autoimmune diseases, and infectious diseases common in individuals with SUD including Hepatitis B and C or HIV/AIDS for the HUD group; 6) MRI contraindications including any metallic implants, pacemaker device, or pregnancy. We did not exclude SUD subjects for history of other drug addiction (e.g., alcohol, marijuana, stimulants/opiates) or other psychiatric disorders with high rates of co-morbidity with these addictions (e.g., depression, post-traumatic stress disorder); 7) healthy control subjects were excluded for a positive breathalyzer test for alcohol or positive urine screen for any psychoactive drugs; and finally, participants were excluded based on 8) MRI quality assurance, including the presence of incidental findings in the WM as indicated by a radiologist, bad diffusion data, or an MRI session that did not include a diffusion sequence (9 CUD/1 HUD/ 7 CTL).

### Behavioral Assessment

In addition to demographic characteristics, neuropsychological tests, estimated verbal [with the reading subtest of the Wide Range Achievement Test-3 (36)], and non-verbal IQ [with the Matrix Reasoning subtest of the Wechsler Abbreviated Scale of intelligence (37)] were assessed in all subjects. Depression was evaluated with the Beck Depression Inventory (BDI) (38). Handedness was assessed with the modified Edinburgh Handedness Inventory (39) (See Table 1).

**Table 1.** Demographic characteristics, neuropsychological measures, and drug-of-choice (DOC) use variables of the study sample.

|  | Healthy<br>Controls<br>N = 58 | Abstinent<br>cocaine users<br>CUD-, N = 32 | Current<br>cocaine users<br>CUD+, N = 28 | MAT<br>heroin users<br>HUD, N = 30 | Statistical analyses |  |
| --- | --- | --- | --- | --- | --- | --- |
|  |  |  |  |  | Test | *p* value |
| *Demographics* |  |  |  |  |  |  |
| Age (years) | 39.3 ± 8.6<sup>a</sup> | 47.3 ± 7.9<sup>b</sup> | 47.7 ± 6.5<sup>b</sup> | 40.3 ± 9.0<sup>a</sup> | F<sub>(3,147)</sub> = 11.0 | **p = .000** |
| Gender (men/women) | 37 / 21 | 28 / 4 | 22 / 6 | 24 / 6 | χ<sup>2</sup> = 7.1 | p = .068 |
| Education (years) | 14.2 ± 2.2<sup>a</sup> | 12.3 ± 1.7<sup>b</sup> | 12.5 ± 1.9<sup>b</sup> | 12.2 ± 2.2<sup>b</sup> | F<sub>(3,147)</sub> = 9.8 | **p = .000** |
| Race (Black or African-American/White/Other & Mixed) | 41<sup>a</sup> / 7<sup>a</sup> / 7 | 21<sup>a</sup> / 4<sup>a</sup> / 4 | 26<sup>a</sup> / 0<sup>a</sup> / 1 | 2<sup>b</sup> / 21<sup>b</sup> / 7 | χ<sup>2</sup> = 64.4 | **p = .000** |
| Intracranial volume (liters) | 1.65 ± .17 | 1.72 ± .14 | 1.65 ± .16 | 1.67 ± .16 | F<sub>(3,147)</sub> = 1.6 | p = .196 |
| *Neuropsychological and self-reported tests* |  |  |  |  |  |  |
| WRAT – Reading Scale (standard score) | 100.5 ± 9.6<sup>a</sup> | 96.2 ± 11.3 | 93.3 ± 11.1<sup>b</sup> | 92.7 ± 11.4<sup>b</sup> | F<sub>(3,147)</sub> = 4.9 | **p = .003** |
| WASI – Matrix Reasoning (scaled score) | 10.5 ± 2.8 | 10.8 ± 2.2 | 9.0 ± 3.2 | 10.6 ± 2.6 | F<sub>(3,145)</sub> = 2.6 | p = .052 |
| Handedness (right/left/ambidextrous) | 54 / 3 / 1 | 26 / 2 / 3 | 27 / 0 / 1 | 26 / 4 / 0 | χ<sup>2</sup> = 9.8 | p = .133 |
| Depression (BDI) | 5.2 ± 7.2<sup>a</sup> | 7.5 ± 8.0<sup>a</sup> | 6.1 ± 6.3<sup>a</sup> | 14.0 ± 12.7<sup>b</sup> | F<sub>(3,143)</sub> = 7.2 | **p = .000** |
| *DOC (and other drug) use variables* |  |  |  |  |  |  |
| Cigarette smokers (Current/Past/Never) | 10<sup>a</sup> / 8 / 37<sup>a</sup> | 19<sup>b</sup> / 9 / 4<sup>b</sup> | 27<sup>c</sup> / 1 / 0<sup>b</sup> | 30<sup>c</sup> / 0 / 0<sup>b</sup> | χ<sup>2</sup> = 91.4 | **p = .000** |
| Fagerstrom Test for Nicotine Dependence | 1.4 ± 1.6<sup>a</sup> | 4.4 ± 2.3<sup>b</sup> | 2.4 ± 1.8<sup>a</sup> | 3.7 ± 1.6<sup>b</sup> | F<sub>(3,80)</sub> = 6.6 | **p = .000** |
| Age of first use (years) | -- | 22.9 ± 6.1 | 20.6 ± 5.9 | 24.6 ± 6.6 | F<sub>(2,88)</sub> = 2.6 | p = .078 |
| Years of regular use | -- | 15.5 ± 8.6<sup>a</sup> | 22.6 ± 8.9<sup>b</sup> | 11.1 ± 7.4<sup>a</sup> | F<sub>(2,89)</sub> = 14.3 | **p = .000** |
| Period of heaviest use (years) | -- | 4.3 ± 3.9 | 7.0 ± 8.2 | 5.7 ± 7.3 | F<sub>(2,86)</sub> = 1.2 | p = .305 |
| Current abstinence (days) | -- | 538.0 ± 1161.9 | 2.8 ± 2.8 | 198.6 ± 265.1 | F<sub>(2,89)</sub> = 4.4 | p = .015 |
| Days of DOC use during past 30 days | -- | 2.3 ± 5.3<sup>a</sup> | 12.1 ± 9.1<sup>b</sup> | 0.2 ± 0.8<sup>a</sup> | F<sub>(2,89)</sub> = 32.3 | **p = .000** |
| Severity of Dependence Scale | -- | 5.6 ± 5.6<sup>a</sup> | 4.5 ± 3.6<sup>a</sup> | 10.2 ± 3.6<sup>b</sup> | F<sub>(2,88)</sub> = 13.5 | **p = .000** |
| DOC withdrawal symptoms (adjusted score) | -- | 14.6 ± 11.5 | 22.8 ± 12.9 | 14.1 ± 14.2 | F<sub>(2,89)</sub> = 4.1 | p = .019 |
| DOC subjective craving (adjusted score) | -- | 8.8 ± 11.5<sup>a</sup> | 24.5 ± 12.4<sup>b</sup> | 16.9 ± 12.5<sup>b</sup> | F<sub>(2,88)</sub> = 16.5 | **p = .000** |

**Notes:** Data expressed as frequencies or means ± standard deviation (SD). In order to correct for multiple comparisons, *p* values were considered significant at *p* < 0.006 (0.05/9 statistical tests) for the demographics/neuropsychological tests and *p* < 0.005 (0.05/10 statistical tests) for the DOC use variables. Post hoc analyses were carried out with Tukey HSD tests for *F* statistics and with Bonferroni-corrected adjusted residuals for chi-square tests. Superscripts refer to the significant between-group post hoc contrasts, e.g., the same superscript means that groups did not differ. Withdrawal symptoms were calculated with two different scales for both DOC, i.e., the Cocaine Selective Severity Scale and the Subjective Opiate Withdrawal Scale. Subjective cravings were also assessed in a drug-specific way, i.e., the 5-items cocaine craving scale and Heroin Craving Questionnaire. HUD withdrawal and craving scores were converted to the CUD scales to allow for statistical comparisons. For 10 of the Table 1 variables (demographics, Neuropsychological tests, and self-reported drug use variables), up to three participants in each group had missing data.

### MRI Acquisition

MRI acquisition was performed using a Siemens 3.0 Tesla Skyra scanner (Siemens Healthineers AG, Erlangen, Germany) with a 32-channel head coil. Diffusion MRI data were acquired using an echo-planar sequence with opposite phase encoding along the left-right axis, monopolar diffusion encoding with 128 diffusion-weighted images (2×64 for each encoding phase) at single shell maximum *b*=1500 s/mm^2^, 13 reference images at b=0 s/mm^2^, field of view (FOV) = 882×1044 mm, 1.8 mm isometric voxel size, repetition time (TR) = 3650 ms, echo time (TE) = 87 ms, bandwidth = 1485 Hz/px, and 80° flip angle, multiband = 3, no in-plane acceleration. A structural T1-weighted scan was acquired using an MPRAGE sequence, sagittal orientation [FOV = 256×256×179 mm^3^; 0.8 mm isotropic resolution; TR 2400 ms; TE 2.07 ms; inversion time 1000 ms; flip angle 8° with binomial (1, −1) fat saturation; bandwidth 240 Hz/pixel; 7.6 ms echo spacing, and in-plane acceleration (GRAPPA) factor of 2] and was used for intracranial volume (ICV) estimation.

### Diffusion Tensor Imaging (DTI) Processing

Diffusion MRI images were preprocessed with the MRtrix3 (40) and FMRIB Software Library (FSL 6.0) (41) toolboxes. Scans were denoised, corrected for eddy current and inhomogeneity distortions and motion artifacts (42) using the “*dwipreproc*” command. All scans were visually inspected to detect major data abnormalities. Diffusion tensors were fitted to each voxel and quantitative maps of diffusion metrics (FA, MD, RD, and AD) were derived from the three orthogonal eigenvectors extracted from each tensor. Fractional Anisotropy reflects the coherence of water diffusion along a specific orientation and MD represents the magnitude of diffusion whereas RD and AD represent the diffusion across the axonal membrane and parallel to the orientation of the axon, respectively. Tract-based spatial statistics (TBSS) was then used to perform whole-brain voxelwise analyses across all participants by, first, aligning and registering individual FA maps to a standard MNI152 template using non-linear registration to create a WM skeleton from the averaged maximal FA (43). Individual DTI maps (of all four metrics) were then projected back onto this skeleton allowing voxelwise group-level statistical tests. In the results section, the extent of the significant voxels is presented as a percentage of the 98,859 voxels comprising the WM skeleton.

### Statistical Analyses

All variables in Table 1 (demographic, neuropsychological, and drug use variables) were compared between healthy controls and all individuals with SUD (CUD−, CUD+, and HUD), or only within SUD as appropriate, using one-way ANOVAs for continuous variables and Chi-square (χ^2^) tests for categorical variables. Post-hoc analyses included Tukey HSD tests for F statistics and Bonferroni-corrected adjusted residuals for Chi-square tests. In order to correct for multiple comparisons, p values for group effects were considered significant at p < .006 (.05/9 tests) for the demographic and neuropsychological measures, and at p < .005 (.05/10 tests) for the drug use variables.

The main set of whole-brain analyses investigated group differences between the healthy controls and individuals with SUD in the four WM diffusion metrics using a design matrix coding for independent groups t-tests (healthy controls vs. SUD). For our second and third objectives, F-statistics maps were computed to identify significant voxels representing the main effect across all four groups (healthy controls, CUD−, CUD+, and HUD). These specific results maps are not presented because of the redundancy with the independent groups t-tests presented in Figure 1. Specific between-subgroups differences were then computed using independent t-statistic contrasts in this main four groups design matrix. All whole-brain WM analyses were carried out with FSL tool “*Randomise*,” a general linear model for non-parametric inferences (43) using 10,000 permutations. To account for multiple comparisons, threshold-free cluster enhancement (TFCE) correction, aiming at better discriminating clustered voxels by enhancing areas of signal exhibiting spatial contiguity, was applied for each analysis (44). A cluster was considered significant when at least 100 contiguous voxels reached the voxelwise threshold of 1-*p*>.949. For our exploratory analyses (aiming at comparing the two CUD subgroups to the HUD group), we reduced the cluster threshold to 15 contiguous significant voxels, and considered anything below our 100 voxels threshold as a trend. Finally, given the expected group differences, we performed correlation analyses to investigate whether any of the observed WM changes were associated with the drug use variables that showed significant differences between the SUD subgroups (years of regular use, days of use in the previous 30 days, severity of dependence, and subjective cravings). Whole-brain voxelwise correlations were performed using z-scored drug use measures across all SUD participants. We estimated the magnitude of the correlations (r values) by averaging the extracted WM metrics from significant voxels and computing the correlations using IBM SPSS statistics version 25 (IBM Corp, Armonk, NY). Because of the exploratory nature of these correlations, significance was considered with a voxelwise threshold of 1-*p*>.949 in at least 100 contiguous voxels. Neuroanatomical localization of WM tracts was done with FSL “*atlasquery”* toolbox and “*JHU ICBM-DTI-81 White-Matter Labels*” atlas (45), with an average probability of region overlap threshold of 2%.

**Figure 1:**
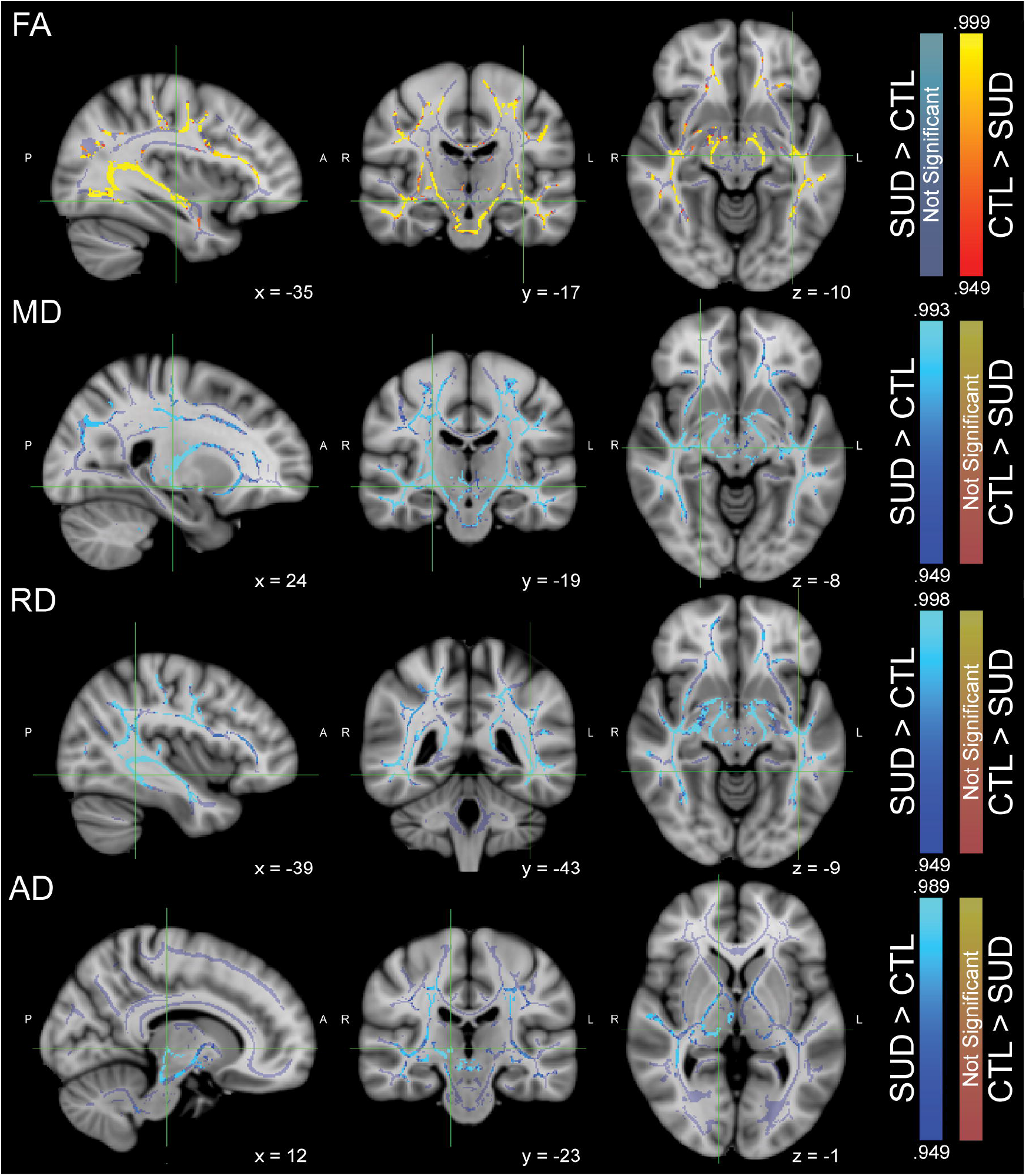
Whole-brain differences between healthy controls (CTL) and individuals with substance use disorder (SUD). This figure represents group difference maps thresholded at significant voxels (1-*p* > .949 corrected). Maps of significant voxels are overlaid on a study-specific white matter skeleton (in light purple) and the MNI152 template. Brain maps are represented according to the radiological convention (the right hemisphere is displayed on the left side). Warm colors represent voxels where healthy controls (CTL) show higher intensity than individuals with SUD whereas cool colors show the opposite. The coordinates of the peak intensity (i.e., the highest p-value) for each image is depicted by a green location cursor.

To control for potential covariates of interest [ICV in addition to demographic and neuropsychological variables that differed between groups: age, education, race, verbal IQ, depression, gender (although the groups did not differ in gender distribution, this variable was included because of its known effect on diffusion metrics (46–48) and because it remains unclear whether there is statistical redundancy while correcting for both gender and ICV (49)), cigarette smoking and nicotine dependence] we performed two sets of analyses. For the continuous covariates, whole-brain correlations were performed with the z-scored covariate of interest; for the categorical variables, independent group analyses (t-tests) were carried out. Due to the number of comparisons (seven variables and four diffusion metrics), the covariate inclusion was determined by a significance threshold of 1-*p*>.9982. None of these analyses reached significance and therefore, with the exception of age and ICV, we did not correct for any of the other variables (that differed between the groups) in the whole-brain group comparisons. No covariates were added to the exploratory correlation analyses.

## Results

### Demographic, neuropsychological, and drug use variables

The groups differed on age, education, verbal IQ, and depression such that healthy controls and HUD subjects were younger than both CUD groups (p<.01), healthy controls had more years of education compared to all SUD groups (p<.01) and higher verbal IQ compared to the CUD+ (p<.05) and HUD subgroups (p<.01), and the HUD group reported higher depression symptoms compared to the other three groups (p<.05) (Table 1). Groups also differed on race, where the HUD group was comprised of significantly more White participants (p<.004) than the other groups. For the drug use variables, groups differed on smoking status where the healthy control group was comprised of significantly more individuals who never smoked (p<.00001), all SUD groups included more current smokers than the control group (p<.003), and the HUD and CUD+ groups included more current cigarette smokers than the CUD−group. The severity of nicotine dependence was significantly higher in the HUD and CUD−groups (p<.05). Significant between-group differences in the primary drug of use (cocaine or heroin) were found for years of regular use, days of drug use during the previous 30 days [higher in the CUD+ group compared to CUD− and HUD (p<.01)], and severity of dependence [higher in the HUD group compared to both CUD groups (p<.001)]. The HUD and CUD+ reported significantly higher subjective craving (p<.01)

### Whole-brain white matter abnormalities in individuals with substance use disorder

The significant whole-brain WM differences between the healthy control subjects and individuals with SUD are presented in Figure 1 with specific clustered results and localizations summarized in Table 2 (Table 5 also summarizes the percentages of significant voxel across the WM skeleton for each contrast/diffusion metric). Individuals with SUD showed significantly lower FA (43.7%, .949<1-*p*<.999) as well as higher MD (56.8%, .949<1-*p*<.993), RD (56.6%, .949<1-*p*<.998), and AD (18.8%, .949<1-*p*<.989) in all major WM tracts when compared to controls. More specifically, individuals with SUD showed lower FA in commissural and projection fibers. Mean diffusivity and RD were significantly higher in the same commissural tracts and projection fibers. Mean diffusivity was also specifically increased in brainstem WM fibers. Finally, individuals with SUD showed higher AD in bilateral projection and in multiple association fibers.

**Table 2.**
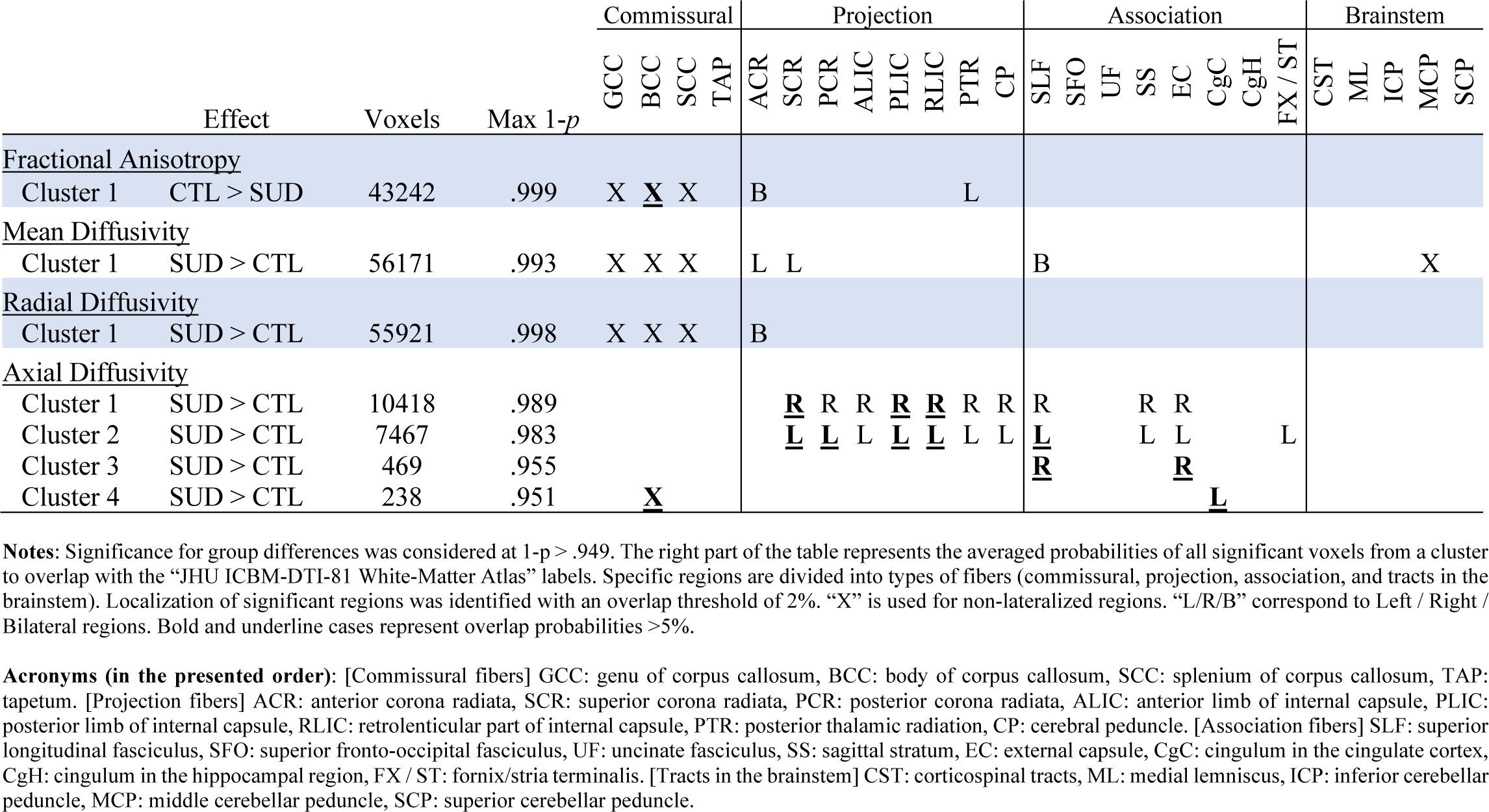
Clustered white matter differences between healthy controls (CTL) and individuals with substance use disorder (SUD)

**Table 3.**
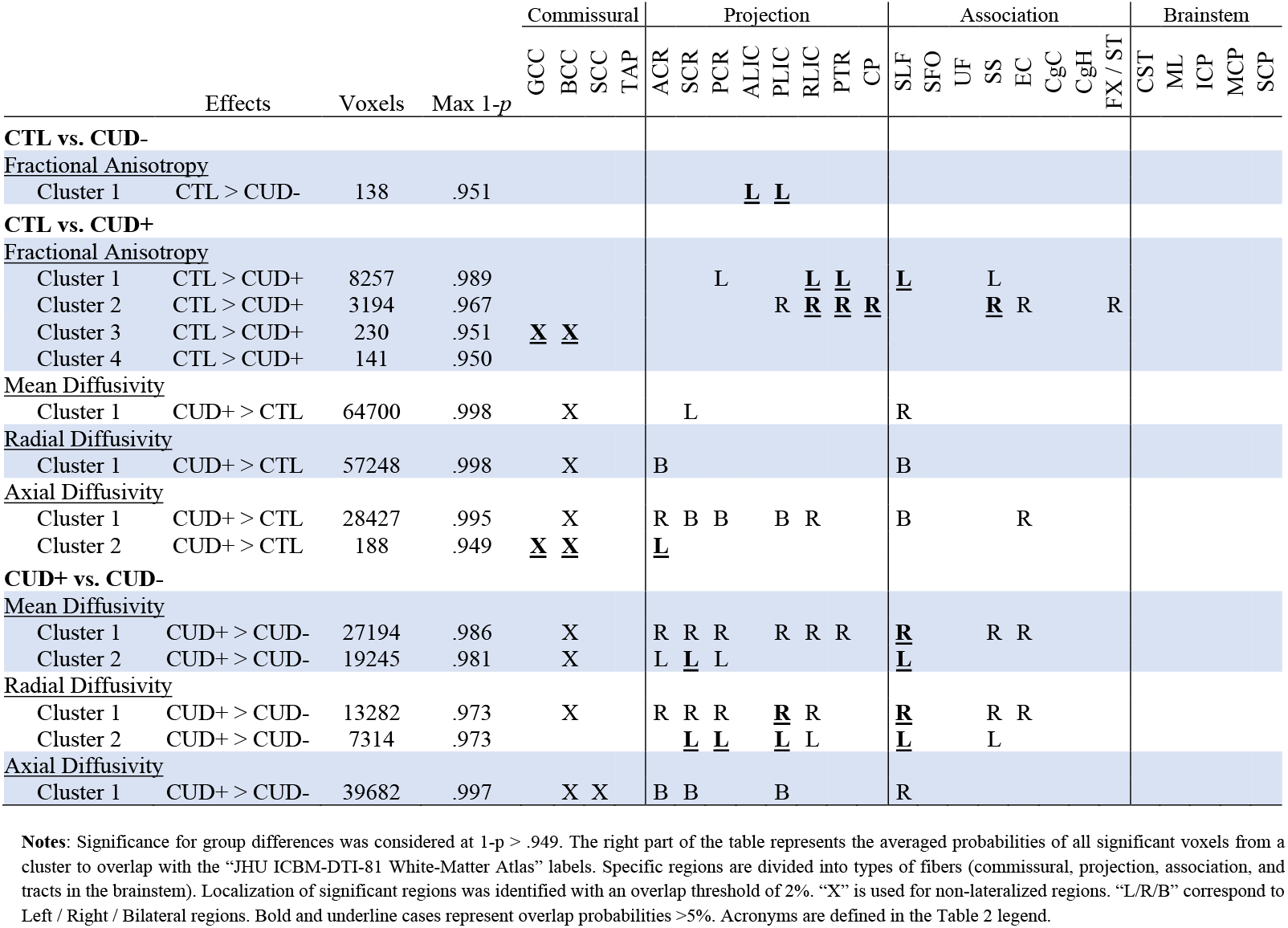
Clustered white matter differences between healthy control subjects (CTL) and abstinent (CUD−) and current cocaine users (CUD+)

**Table 4.**
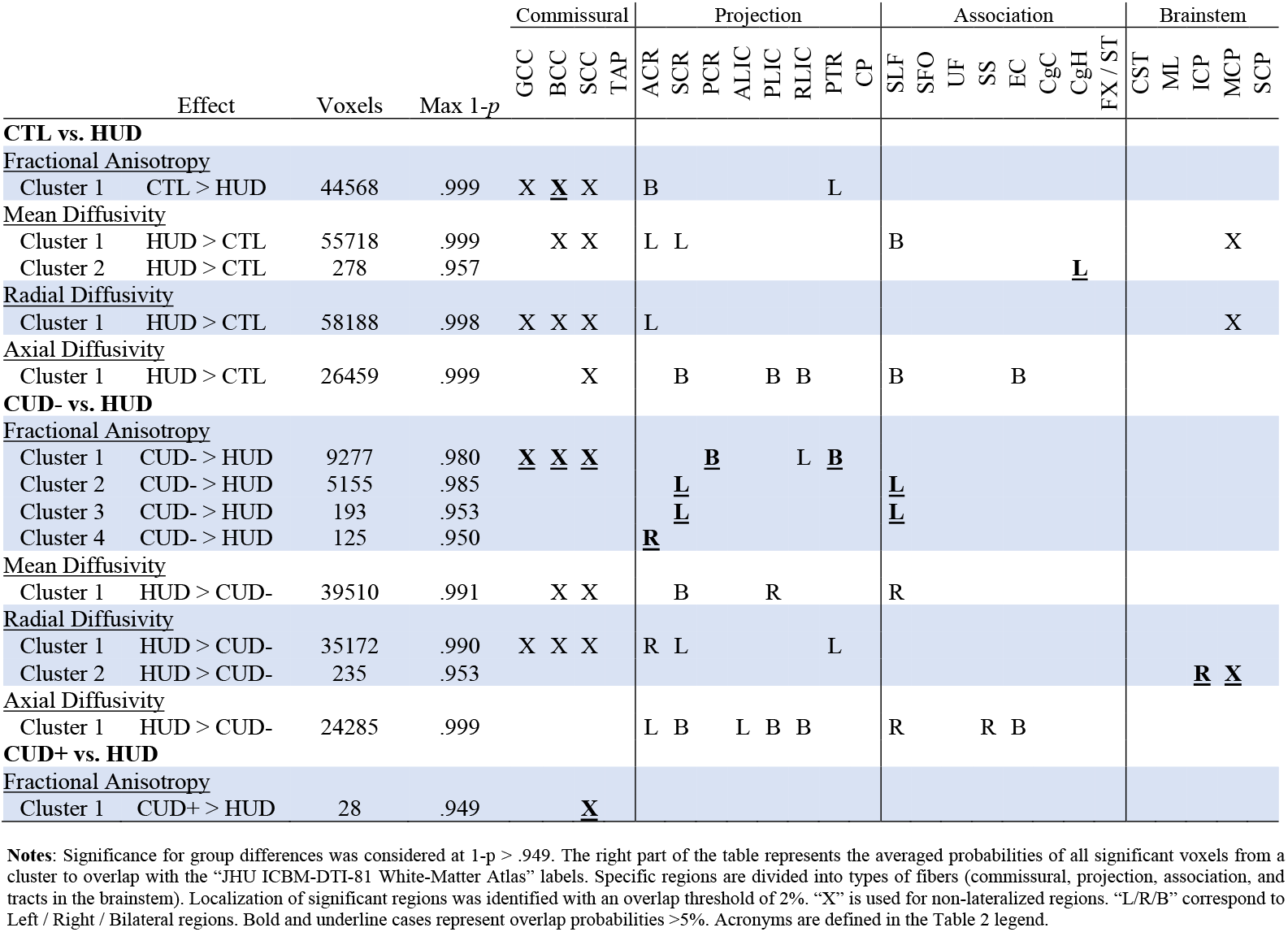
Clustered white matter differences between healthy control subjects (CTL), abstinent (CUD−) and current individuals with cocaine (CUD+), and individuals with heroin use disorder (HUD)

**Table 5.** Summary of percentages of significant voxels in the WM skeleton for each contrast.

| Contrasts | Fractional Anisotropy | Mean Diffusivity | Radial Diffusivity | Axial Diffusivity |
| --- | --- | --- | --- | --- |
| CTL > SUD | 43.7% | -- | -- | -- |
| SUD > CTL | -- | 56.8% | 56.6% | 18.8% |
| CTL > CUD- | .17% | -- | -- | -- |
| CUD- > CTL | -- | -- | -- | -- |
| CTL > CUD+ | 11.9% | -- | -- | -- |
| CUD+ > CTL | -- | 65.5% | 57.9% | 28.9% |
| CTL > HUD | 45.1% | -- | -- | -- |
| HUD > CTL | -- | 56.6% | 58.9% | 26.8% |
| CUD- > CUD+ | -- | -- | -- | -- |
| CUD+ > CUD- | -- | 46.1% | 20.9% | 40.1% |
| CUD- > HUD | 15.0% | -- | -- | -- |
| HUD > CUD- | -- | 40.0% | 35.8% | 24.6% |
| CUD+ > HUD | .03% | -- | -- | -- |
| HUD > CUD+ | -- | -- | -- | -- |

### Group-specific whole-brain abnormalities in abstinent and current cocaine users

For this section and the one below (comparisons with HUD), all significant contrasts are presented in Figure 2 for the FA and MD results and in Figure 3 for the AD and RD results. Clustered results with corresponding atlas location are also presented in Table 3 (see also Table 5). Control subjects showed higher FA than the CUD−group in a small cluster located in left projection fibers (.17%, .949<1-*p*<.951). No other diffusion metric showed significant effect for this contrast. Global WM abnormalities were also found when comparing control subjects to the CUD+ group encompassing decreased FA (11.9%, .949<1-*p*<.989), as well as increased MD (65.5%, .949<1-*p*<.998), RD (57.9%, 0.949<1-*p*<0.998), and AD (28.9%, .949<1-*p*<.995). More specifically, decreased FA was found in commissural, projection, and in association fibers. The other diffusion metrics were increased in a significant portion of the WM skeleton similarly encompassing commissural, projection, and association fibers. Finally, when directly compared with CUD−, the CUD+ group showed higher MD (46.1%, .949<1-*p*<.986), RD (20.9%, .949<1-*p*<.973), and AD (40.1%, .949<1-*p*<.997) in a substantial part of the WM skeleton including commissural fibers, most projection fibers as well as association fibers.

**Figure 2:**
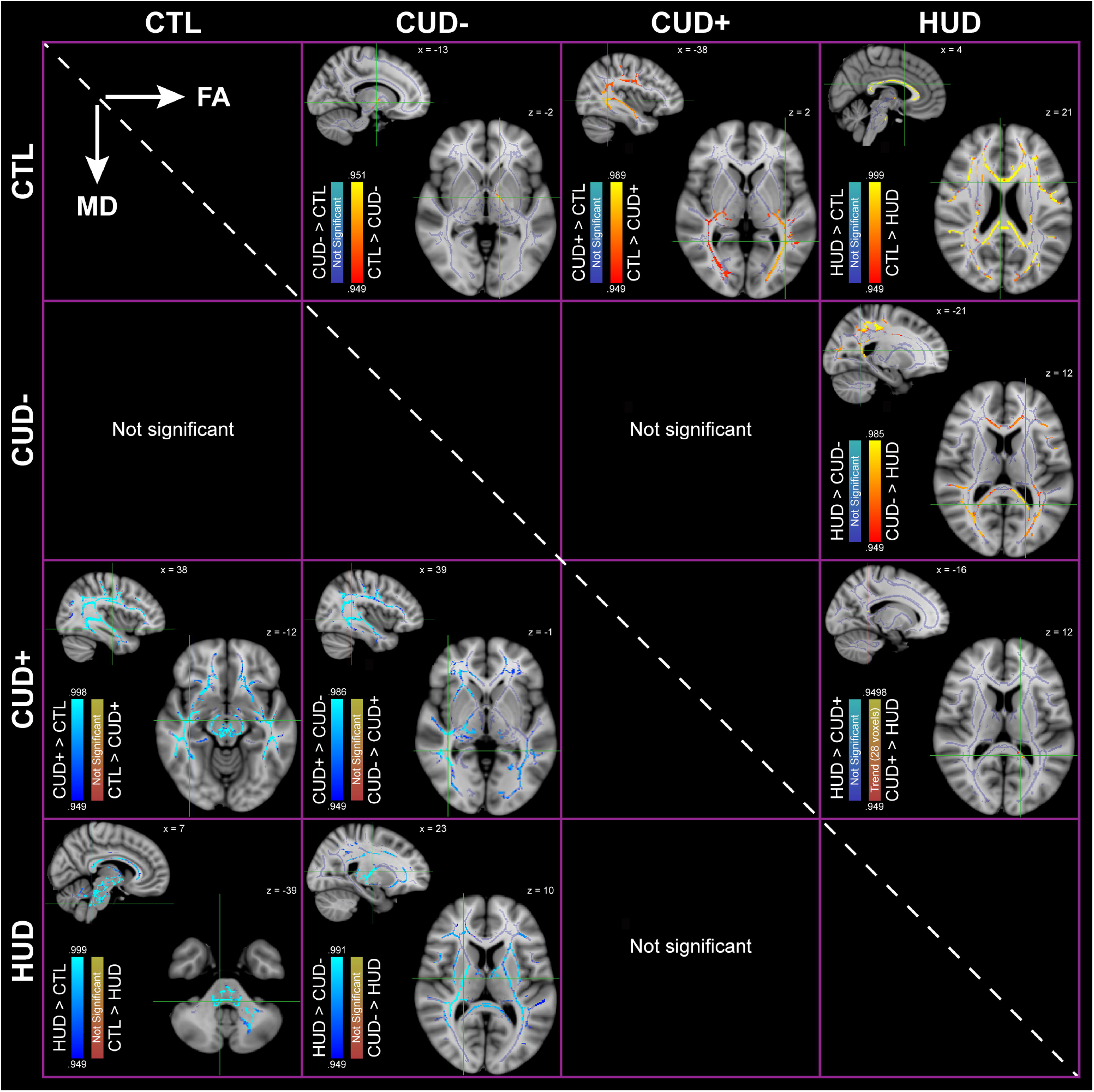
Group-specific whole-brain differences of fractional anitotropy (FA) and mean diffusivities (MD). This figure represents thresholded maps of significant voxels (1-p > .949 corrected) between all study groups [healthy controls (CTL), abstinent cocaine users (CUD−), current cocaine users (CUD+), and medically-assisted treatment heroin users (HUD)] on FA and MD. Legends show the direction of the effects for each case. Maps of significant voxels are overlaid on a study-specific white matter skeleton (in light purple) and the MNI152 template. Brain maps are represented according to the radiological convention (the right hemisphere is displayed on the left side). The coordinates of the peak intensity (i.e., the highest p-value) for each image is depicted by a green location cursor.

**Figure 3:**
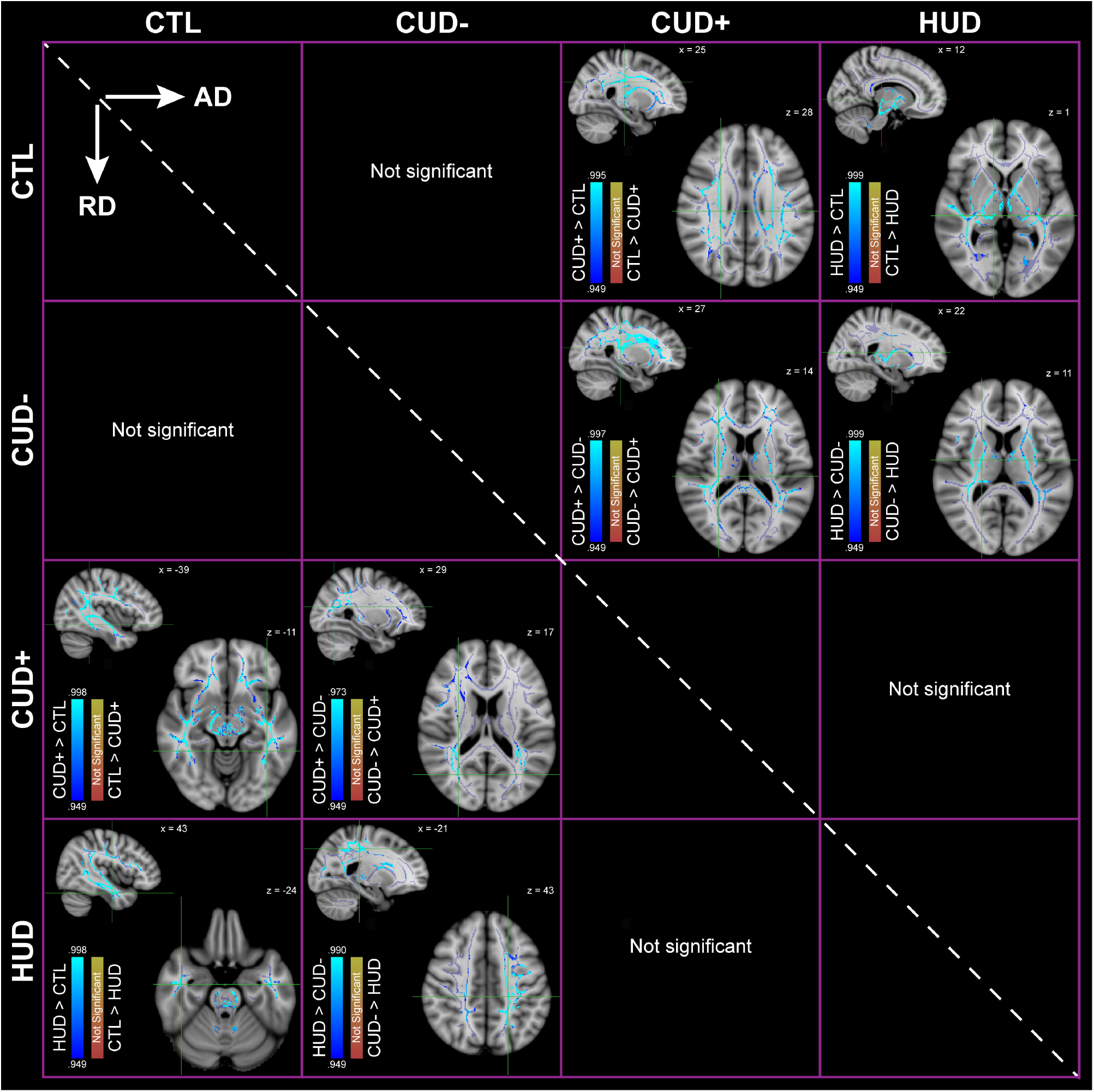
Group-specific whole-brain differences of axial (AD) and radial diffusivities (RD). This figure represents thresholded maps of significant voxels (1-*p* > .949 corrected) between all study groups [healthy controls (CTL), abstinent cocaine users (CUD−), current cocaine users (CUD+), and medically-assisted treatment heroin users (HUD)] on AD and RD. Legends show the direction of the effects for each case. Maps of significant voxels are overlaid on a study-specific white matter skeleton (in light purple) and the MNI152 template. Brain maps are represented according to the radiological convention (the right hemisphere is displayed on the left side). The coordinates of the peak intensity (i.e., the highest p-value) for each image is depicted by a green location cursor.

### Group-specific whole-brain abnormalities in cocaine and heroin use disorder

We also explored whether the whole-brain WM abnormalities observed in the CUD groups were generalizable to individuals with HUD. In addition to Figures 2–3, clustered results are presented in Table 4 (see also Table 5). When compared to healthy controls, individuals with HUD showed a general decrease in FA and increased MD, RD and AD (respectively 45.1%, 56.6%, 58.9%, and 26.8%, all .949< 1-*p* <.999). Specifically, FA changes were found in commissural and projection fibers, while the MD, RD, and AD effects were similarly found in commissural and projections fibers, but also in association fibers (for MD and AD) and fibers emanating from the brainstem (for MD and RD). Similar effects were found when comparing individuals with HUD to CUD−[decreased FA (15.0%, .949<1-*p*<.985) and increased MD (40.0%, .949<1-*p*<.991), RD (35.8%, .949<1-*p*<.990), and AD (24.6%, .949<1-*p*<.999)]. This decreased FA was found in commissural, projection, and association fibers. Increased MD, RD, and AD were also found in commissural, projection, association fibers (for MD and AD only), as well as in fibers connecting the brainstem (for RD only). Interestingly, a trend for a significant reduction in FA was also observed when comparing individuals with HUD to those with CUD+ in part of the splenium of the corpus callosum (28 voxels, .03%, 1-*p*= .949).

### Correlations between drug use variables and white matter diffusion metrics

Across all SUD, whole-brain voxelwise correlation analyses showed significant negative correlations between regular use and FA (.67%, .949<1-*p*<.974, extracted cluster: r=.49, p<.00001) and AD (5.1%, .949<1-*p*<.965, extracted cluster: r=.42, p<.0001) where more years of regular use was associated with decreased FA and AD (Figure 4A). Significant positive correlations between subjective craving scores and WM diffusivities were also found (Figure 4B) showing that higher baseline craving was associated with higher MD (48.1%, .949<1-*p*<.989, extracted cluster: r=.34, p<.01), RD (11.2%, .949<1-*p*<.971, extracted cluster: r=.42, p<.0001), and AD (33.5%, .949<1-*p*<.992, extracted cluster: r=.46, p<.00001).

**Figure 4:**
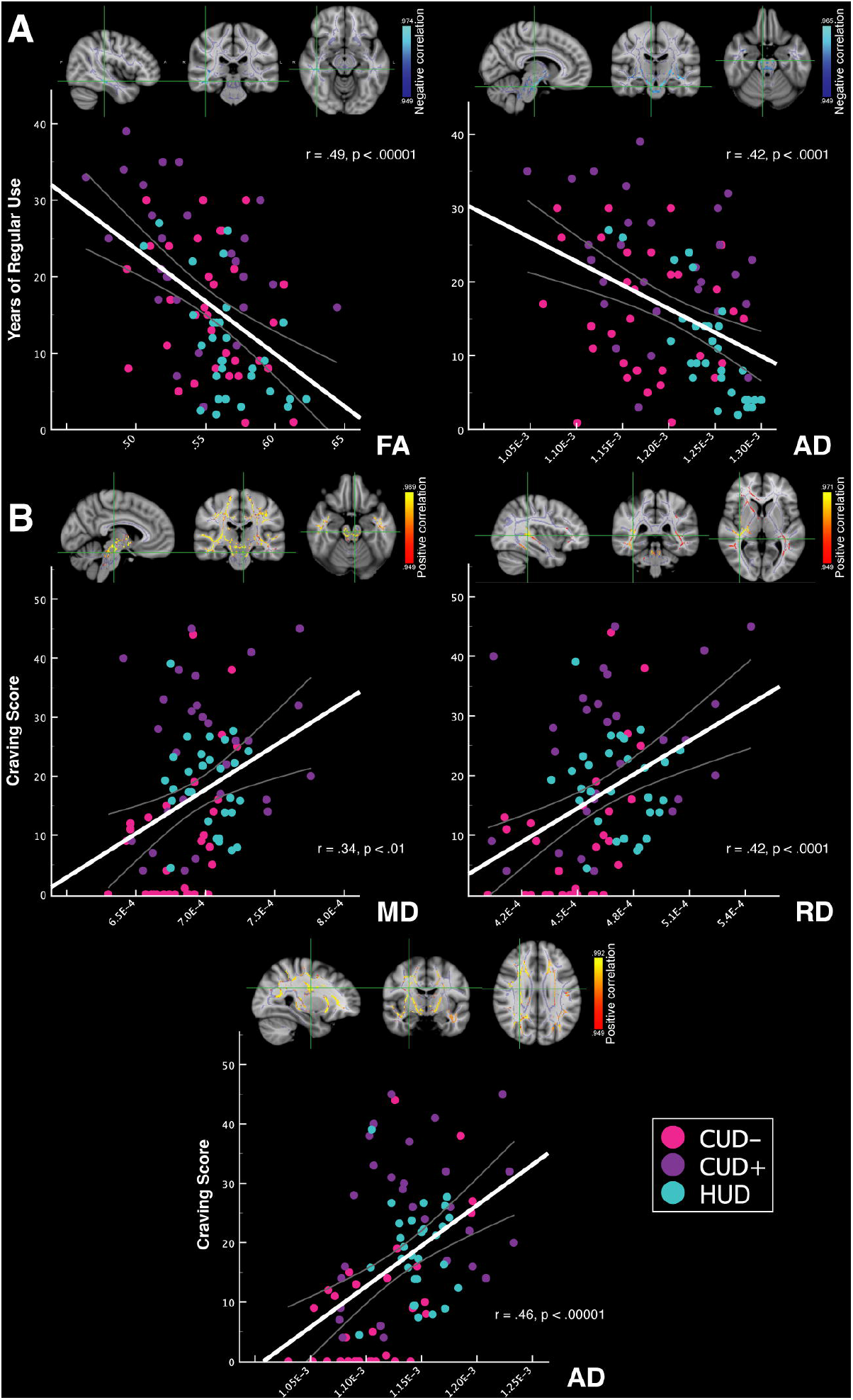
Significant whole-brain voxelwise correlations between drug use variables and WM diffusion metrics in individuals with SUD. This figure shows scatterplots of the correlations between years of regular use (A) and craving scores (B) with the averaged extracted diffusivity values across significant (1-p > .949 corrected) voxels for the fractional anisotropy (FA), mean (MD), radial (RD), and axial diffusivities (AD) in abstinent cocaine users (CUD−, pink), current cocaine users (CUD+, purple), and medically-assisted treatment opiate users (HUD, cyan). The regression line (white) represents the correlation across the entire SUD group. Maps of significant voxels are overlaid on a study-specific white matter skeleton (in light purple) and the MNI152 template. Brain maps are represented according to the radiological convention (the right hemisphere is displayed on the left side). The coordinates of the peak intensity (i.e., the highest p-value) for each image is depicted by a green location cursor.

## Discussion

The goal of this study was to assess WM abnormalities in three groups of drug-addicted individuals to explore the generalized vs. acute/specific drug effects using highly reliable acquisition parameters and analytical specificity. Our study replicated previous reports in the literature, consistent with an emerging consensus supporting widespread WM microstructure abnormalities, characterized by decreased FA, and increased MD, RD, and AD, across all major WM tracts, generalizable across individuals with SUD. Our results showed that between 19% and 57% of the WM skeleton was affected. In addition to the comparison between psychostimulants and opiates for generalizability purposes, we also investigated the contribution of recency of drug use (in the CUD+ and CUD−, who used cocaine on average within three days or 18 months of the study, respectively, and in the HUD, all currently using opiates as part of MAT). Results showed a more detrimental effect of recency of drug use (most severity in the HUD and CUD+ vs. CUD−) and more severity in heroin vs. cocaine users (most severity in the HUD vs. CUD). Among all individuals with SUD, longer periods of regular use and higher subjective cravings were associated with decreased FA and increased MD, RD, and AD.

Individuals with HUD showed the most pronounced impairments followed by the CUD+, with the CUD−showing the least impairment (different from healthy controls only in a small cluster located in projection fibers of the internal capsule). Specifically, the HUD group showed more widespread FA impairments than CUD+ (45.1% vs. 11.9% of WM skeleton); a direct comparison revealed a significant difference limited to the splenium of the corpus callosum (higher FA in CUD+ > HUD). This pattern of results, and correlations with craving (the higher the craving, the higher the MD, RD, and AD across all SUD), suggest the detrimental effects of acute/recent drug use on the brain (indicated by a positive drug urine status for both CUD+ and HUD groups), with potentially more specificity and severity of the effect for opiates vs. stimulants. However, severity of dependence (highest in the HUD group, all inpatients on MAT) and more years of regular use (highest in the CUD+) may have also contributed to this pattern of results. Indeed, across all individuals with SUD, more years of regular use was associated with a reduction of FA and AD. Similar associations between WM abnormalities with duration of regular use were previously reported in the tapetum in CUD (10) and in frontal pathways in HUD (14,17) [and in the inferior frontal gyrus in alcohol use disorder (50)]. Of note are the seemingly contradictory results for AD (i.e., decrease with years of use and increase with craving). Upon closer inspection, there was a different localization with minimal overlap (<3% of the WM skeleton) of the maximal peak, with a specific subcortical involvement for the association with regular use and a more widespread cortical effect for the association with craving. Taken together, these results support the detrimental effects of both chronicity and acuteness of use of both drug types.

The white matter abnormalities across both substance types documented in our study are consistent with previous diffusion MRI findings in cocaine (5,6,8–11) and heroin users (12–17) as well as in other types of drug users (4,50–56) when compared to demographically matched healthy controls. However, our results highlight a more extensive pattern of WM abnormalities than previously reported, encompassing fronto-striatal and fronto-temporal projections, involved in the regulation of learning and memory, executive control, and reward-driven behaviors, but also centro-parietal projections and association WM tracts (such as the corona radiata, the posterior thalamic radiation, and the superior longitudinal fasciculus), which constitute major intrahemispheric connections. In general, the pattern observed in the SUD subjects (reduced FA concomitantly with increases in MD and RD but also AD) is indicative of WM impairment, as has been shown in brain disorders with both normal-appearing or abnormal WM including multiple sclerosis, stroke, dementia, and schizophrenia (57–59). These changes in WM have been associated with neurobiological correlates of demyelination, decreased axonal packing (60,61), axonal degeneration, axonal loss (62,63), neurofilament damage and WM fiber atrophy, and increased extracellular water content (61,64,65), although these effects remain to be validated by preclinical/ex vivo studies.

Importantly, our results confirmed the generalization of these WM impairments across two different classes of drugs of abuse (stimulants and opiates) that are known for their neurotoxic effects associated with neuroinflammatory brain responses (66,67), which may in turn precipitate myelin degradation (68,69). Shared neurobiological mechanisms contributing to these widespread WM insults may involve oxidative stress responses, down-regulation of myelin-related protein expression, mitochondrial dysfunction, and/or neuronal apoptosis (11,70–73). Other processes may also include reduced efficiency of glial cells in regulating glutamate homeostasis and blood-brain barrier dysfunctions exposing the brain to toxins (67,74). Axonal and myelin degeneration could also result from vascular effects of both classes of drugs through stimulant-induced vasoconstriction, increasing the risk of hypoperfusion (75,76), and/or through opiate-related ischemic lesions and perfusion deficits, potentially induced by respiratory suppression, altered consciousness, overdoses, or other vascular conditions (e.g., vasculitis and rhabdomyolysis) that are often observed in heroin/opiate users (77,78).

Beyond the precise neurobiological mechanisms, the importance of these results is in their potential for outcome prediction: in other studies of individuals with HUD undergoing MAT, these WM abnormalities were observed in those who relapsed (positive urine status for both methadone and heroin) as compared to those who abstained (positive urine status for methadone but not heroin); further recovery was associated with less severe impairments in individuals with HUD in prolonged abstinence (negative urine status) (13,79,80). Positive correlations between long-term abstinence and WM integrity in the ventromedial PFC/orbitofrontal cortex in individuals with CUD and in frontal WM in opiate/heroin users (8,12,17) also support this possibility. Our own results in the CUD−similarly suggest that longer abstinence length durations (and/or less chronicity of use) could potentially contribute to WM recovery as remains to be tested in longitudinal studies. Nevertheless, it is important to consider that some of the WM microstructure impairments may be persistent, as suggested by the significant FA reductions in this group compared to healthy controls, consistent with similar results in the internal capsule of individuals with methamphetamine use disorder (81).

Several limitations are important to consider. First, longitudinal within-subject studies are needed to ascertain the impact of abstinence, and other time-varying metrics, on our results. Further, in addition to the three SUD groups included here, including groups of current heroin users and those entirely abstinent (inclusive of MAT, HUD-) would help differentiate between the acute effect of the drug and the specificity of impairments to stimulants vs. opiates. Future studies should also include polysubstance users, as increases in demyelination in PFC regions (i.e., increased RD) were previously associated with the number of substances used (82), potentially indicative of a dose effect. In addition, more women should be included to allow the study of possible sex differences in WM microstructure effects, as suggested in a population study of WM integrity (83). Future studies should also aim at assessing the integrity of specific WM tracts by using advanced diffusion methodology (i.e., orientation distribution function) to model specific components of the salience/reward, executive control, and learning networks, and explore associations between these WM abnormalities with impaired behavioral and neurocognitive functioning in SUD.

In conclusion, we report whole-brain widespread WM abnormalities across individuals with SUD as driven by HUD on MAT and CUD+, and correlations with craving that together suggest the impact of the acute effects of both opiates and stimulants on WM integrity, with results also pointing to the specificity of this effect (i.e., reduced FA for HUD>CUD+ in the splenium of the corpus callosum). The lower extent of impairment in the CUD−corroborates previous findings of brain recovery with abstinence/less current use but the persistence of these impairments, and correlations with years of regular use across all SUD, also suggest the detrimental effect of chronicity of drug use on WM integrity in drug addiction.

## Data Availability

All data produced in the present study are available upon reasonable request to the authors.

## Acknowledgments

This work was supported by the National Institute on Drug Abuse (Goldstein, 1R01DA048301-01A1), the National Center for Complementary and Integrative Health (Goldstein, 1R01AT010627-01), and the Canadian Institutes of Health Research (Gaudreault, postdoctoral research fellowship).

